# Oral diadochokinesis, tongue pressure, and lip-seal strength in Japanese workers: A cross-sectional study

**DOI:** 10.1101/2024.06.05.24308513

**Authors:** Akira Minoura, Yoshiaki Ihara, Hirotaka Kato, Kouzou Murakami, Yoshio Watanabe, Kojiro Hirano, Yoshinori Ito, Akatsuki Kokaze

## Abstract

This cross-sectional study investigated the correlation between lip-seal strength, tongue pressure, and oral diadochokinesis (OD) in Japanese workers. The relationships between lip-seal strength, tongue pressure, and OD by age groups were investigated using multiple regression analyses on 496 workers. OD was measured using the number of consecutive “Pa,” “Ta,” and “Ka” vocalizations that could be produced in 5 s. In this study, 478 participants (437 males and 41 females) were included in the analysis after excluding 18 participants who could not complete all oral cavity measurements. This study revealed a significant positive correlation between “Pa,” “Ta,” and “Ka,” with correlation coefficients of 0.500–0.665. Lip-seal strength only significantly correlated positively with “Pa”; however, tongue pressure significantly correlated positively with all of “Pa,” “Ta,” and “Ka” vocalizations. Regarding body mass index (BMI), no significant relationship with either “Pa,” “Ta,” or “Ka” was observed. As a result of aging, “Ta” and “Ka” showed a significant negative correlation with age. Multiple regression analyses, which included age, sex, BMI, alcohol consumption, and smoking, revealed a strong association between “Pa,” “Ta,” and “Ka” and lip-seal strength. However, only “Pa” showed a significant correlation with tongue pressure. Even in young and middle-aged adults, OD may be associated with lip-seal strength and tongue pressure. Measuring OD through dental screening of workers will help prevent the disease across a wide age range.

## Introduction

Oral diadochokinesis (OD) is a test that evaluates the mobility of the oral organs, including the tongue, lips, and jaw, particularly among the elderly [1, 2]. It uses repetitive vocalizations of vowels and consonants to measure the accuracy, speed, and consistency of oral movements [3]. Dentists or speech-language pathologists primarily perform OD to evaluate movement disorders caused by dysarthria, delayed speech development, and neurological disorders [4]. In contrast, in several neurological disorders, tongue pressure and lip-seal strength may be sensitive indicators of articulatory function and oral motor impairment [5]. Although tongue pressure, lip-seal strength, and OD may be associated with diseases in the elderly, how these factors relate to OD in younger and middle-aged individuals remains unclear[1, 4]. We have shown the trend of tongue pressure and lip-seal strength by age group among Japanese workers [5]. In particular, lip-seal strength decreased in young individuals compared with that in older adults, and tongue pressure decreased in young individuals compared with that in middle-aged adults [5]. Measuring tongue pressure and lip-seal strength requires a special instrument; however, OD does not require special equipment and is easy to measure; therefore, investigating the relationship between tongue pressure, lip-seal strength, and OD can lead to a simple measurement of the oral environment or prevent oral disorders[4, 6]. Enhancing working conditions may prevent daytime sleepiness due to obstructive sleep apnea, according to previous research [7, 8].

Building on these findings, this study suggests that improving OD increases tongue pressure and lip-seal strength, ultimately reducing daytime sleepiness among workers. Increasing OD could help reduce sleep-related accidents among workers, particularly the elderly, at a cheap cost because the Japanese government is encouraging the employment of the elderly, whose percentage is rapidly rising in the country [7]. Because obstructive sleep apnea is believed to be intensified by age-related tongue root depression, OD improvement may help avoid daytime sleepiness [10]. Particularly in jobs requiring life-threatening duties, including security guards and drivers, measuring OD is crucial to help forecast excessive daytime sleepiness. Therefore, based on earlier studies, we postulated that OD, tongue pressure, and lip-seal strength are related and may vary with age.

Consequentially, this study aimed to examine the correlation among tongue pressure, lip-seal strength, and OD across various age groups, with the aim of averting such disorders. We investigated the correlation between tongue pressure, lip-seal strength, and OD in Japanese workers, categorized by age group.

## Materials and Methods

### Study design

This study, which was conducted between November 2021 and June 2022, involved 496 employees (454 males and 42 females) from two Japanese taxi companies (the recruitment period: from November 9 2021 to June 22 2022). After excluding 18 participants who could not complete all oral cavity measurements, the analysis included 478 participants (437 males and 41 females). Before the study began, the Medical Ethics Committee of Showa University School of Medicine (21-088-A) approved the survey and collected written informed consent from each participant.

### Measurements

An experienced dentist instructed the study participants in the OD task measurement and recorded the results. We instructed the participants to repeat a given syllable, “Pa,” “Ta,” and “Ka,” as fast as possible on one breath in 5 s [4]. Furthermore, we trained a dentist in the measurement of tongue pressure and lip-seal strength among all participants. We used a reliable specialized device (Lipple Kun, Shofu, Kyoto, Japan) to measure the lip-seal strength [11, 12-14]. We measured tongue pressure using specialized equipment (TPM-01; JMS Co. Ltd., Hiroshima, Japan). We determined the participants’ tongue pressure by averaging the three recordings. We took each measurement after a minimum of 1 min of rest following the previous measurement [8].

### Covariates

We used a self-administered questionnaire with items on age, alcohol use (once a week or more, less than once a week), smoking habits (yes, no), weight (kg), and height (cm) because studies have associated alcohol use, smoking, and overweight/obesity with tongue pressure and lip-seal strength among healthy Japanese individuals [15]. The COVID-19 pandemic in Japan has led to changes in job descriptions, such as from drivers to clerks, which we did not include in the questionnaire.

### Statistical analysis

We used the Shapiro–Wilk test for continuous factors to verify a normal contribution. This study displays median values, specifically the 25th and 75th percentiles, for continuous variables with non-normal distribution. We computed correlation coefficients (Pearson’s r) before performing the analysis to verify multicollinearity between tongue pressure, OD, lip-seal strength, and other variables. *P*-values < 0.05 were used to denote statistical significance. In the primary statistical analysis, we investigated the age-group-specific relationships between tongue pressure, lip-seal strength, and OD using multiple regression analysis. We have presented the data along with 95% confidence intervals. This study adhered to the Strengthening the Reporting of Observational Studies in Epidemiology standard for conducting cross-sectional studies. Statistical analyses were performed using JMP (version 17.0; SAS Institute, Inc., Cary, NC, USA).

## Results

In this study, to examine the characteristics of OD by age, we stratified the 478 subjects into those in their 20s, 30s, 40s, 50s, and >60s. This study excluded participants under the age of 20 years. Table 1 shows the attributes of the subjects categorized by age group.

**Table 1.** Characteristics of the study subjects by age group.

|  | **Total** | **20s** | **30s** | **40s** | **50s** | **60s and over** | <sup>1)</sup>**p-value** |
| --- | --- | --- | --- | --- | --- | --- | --- |
|  | (N = 478) | (n = 111) | (n = 65) | (n = 90) | (n = 151) | (n = 61) |  |
| **Lip-seal strength** | 13.5 (11.4, 16.2) | 11.6 (9.5, 13.7) | 12.9 (11.4, 15.7)* | 14.6 (12.3, 17.1)* | 14.3 (12.2, 16.9)* | 14.9 (11.8, 17.9)* | <0.001 |
| **Tongue pressure** | 41.7 (35.5, 48.4) | 39.8 (33.3, 46.1) | 44.4 (37.7, 50.0)* | 44.3 (35.3, 50.1)* | 42.0 (36.7, 47.2) | 41.0 (34.6, 46.4) | 0.012 |
| **Age** | 47.0 (31.0, 55.0) |  |  |  |  |  |  |
| **Female** | 41 (8.6) | 26 (23.4) | 2 (3.1) | 3 (3.3) | 8 (5.3) | 2 (3.3) | <0.001 |
| **BMI** | 24.1 (21.3, 27.1) | 21.7 (19.7, 24.5) | 24.1 (22.3, 28.2)* | 24.6 (22.3, 27.5)* | 24.5 (22.2, 27.4)* | 25.4 (23.1, 28.7)* | <0.001 |
| **Pa** | 33 (31, 35) | 33 (32, 35) | 33 (30, 36) | 34 (32, 36) | 33 (31, 34) | 32 (29, 33)* | <0.001 |
| **Ta** | 35 (32, 38) | 36 (33, 39) | 35 (33, 40) | 36 (33, 39) | 34 (32, 37) * | 32 (30, 34)* | <0.001 |
| **Ka** | 32 (29, 34) | 33 (30, 35) | 32 (30, 36) | 33 (29, 36) | 31 (29, 33) | 29 (26, 32)* | <0.001 |
| **Alcohol use** | 275 (57.5) | 55 (49.6) | 35 (53.9) | 59 (65.6) | 84 (55.6) | 42 (68.9) | 0.06 |
| **Smoking** | 169 (37.2) | 29 (26.1) | 23 (35.4) | 44 (48.9) | 53 (35.1) | 20 (32.8) | 0.021 |

Except where indicated n (%), values are median (25th, 75th percentile). BMI: Body mass index. <sup>1)</sup>Analysis of variance was performed for continuous variables, 113 and Chi-square test was performed for dichotomous variables. \* The results of the Dunnett's test by age, using the 20s as controls, were significantly lower or 114 higher. 115

Table 2 shows the correlation coefficients between OD and covariates, excluding workers aged ≥60 years. The results indicate a significant positive correlation between “Pa,” “Ta,” and “Ka,” with correlation coefficients of 0.500–0.665. Tongue pressure had a noteworthy positive correlation with “Pa,” “Ta,” and “Ka,” although lip-seal strength only displayed a significant positive correlation with “Pa.” No significant relationship was observed between BMI and “Pa,” “Ta,” and “Ka.” Furthermore, “Ta” and “Ka” showed a significant negative correlation with age. Upon evaluation, we determined that none of the factors strongly correlated with any of the other covariates. To prevent multicollinearity, we decided to incorporate all factors into the analysis

**Table 2.** Correlation coefficients between oral diadochokibesis and covariates (excluded 60s and over)

|  |  |  |  | Pearson's r |  |  |  |
| --- | --- | --- | --- | --- | --- | --- | --- |
|  | **a** | **b** | **c** | **d** | **e** | **f** | **g** |
| **a. Pa** |  | 0.606 * | 0.500 * | 0.121 * | 0.184 * | −0.016 | −0.021 |
| **b. Ta** |  |  | 0.665 * | 0.207 | 0.202 * | −0.098 * | 0.020 |
| **c. Ka** |  |  |  | 0.446 | 0.144 * | −0.101 * | −0.037 |
| **d. Lip-seal strength** |  |  |  |  | 0.401 * | 0.321 * | 0.307 * |
| **e. Tongue pressure** |  |  |  |  |  | 0.119 * | 0.292 * |
| **f. Age** |  |  |  |  |  |  | 0.210 * |
| **g. BMI** |  |  |  |  |  |  |  |

Values are correlation coefficient. BMI: Body mass index. \*p < 0.05.

Table 3 presents a concise overview of the relationship between lip-seal strength, tongue pressure, and OD across different age groups. We conducted several regression models to separately investigate the effects of lip-seal strength and tongue pressure on OD. Model 1 found a significant correlation between “Pa,” “Ta,” and “Ka” and tongue pressure. Model 2 found a significant correlation between “Pa” and lip-seal strength. We adjusted both models for sex, age, BMI, alcohol consumption, and smoking.

**Table 3.**
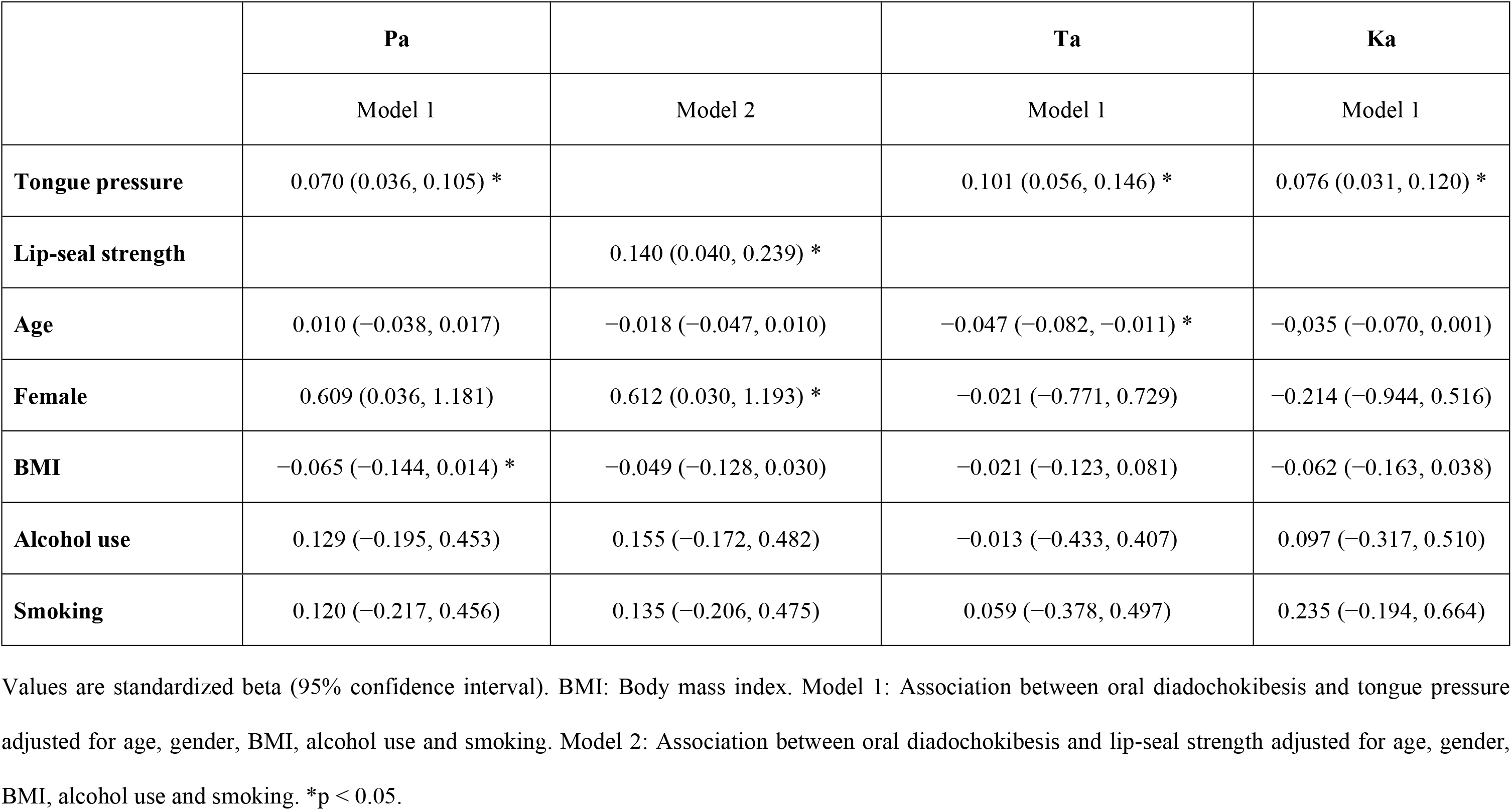
Associations between tongue pressure, lip-seal strength, and oral diadochokibesis (Results of multiple regression analyses)

## Discussion

This appears to be the first investigation into the correlation between OD in Japanese individuals and specific lip-seal strength and tongue pressure. After adjusting for age, sex, BMI, alcohol consumption, and smoking, the results showed a positive correlation between tongue pressure and “Pa,” “Ta,” and “Ka.” Moreover, lip-seal strength showed a positive correlation with “Pa.” Regardless of age, these results showed that good OD can help maintain lip-seal strength and tongue pressure, which may help prevent oral disorders due to deterioration with age [4, 8]. According to the results of this study, OD has mostly been used to evaluate frailty and oral diseases in older individuals, at least in Japanese studies before this one [1]. However, it may also be used to determine health-related outcomes in younger groups [1, 9, 10]. Moreover, recent studies have shown that daytime sleepiness is a serious risk factor for insulin resistance [11]. Regardless of age or weight, daytime sleepiness may contribute to the development of insulin resistance [12]. OD may also contribute to the association between daytime sleepiness and diabetes mellitus and risk factors for obesity and aging. Future research should examine causal relationships among eating patterns, daytime sleepiness, diabetes mellitus, and oral environment markers, including lip-seal strength and tongue pressure. Training perioral muscle function may improve percutaneous oxygen saturation during sleep, despite reports linking it to sleep disturbances and diabetes mellitus [20]. Interventions can help modify eating and OD behaviors, which could prevent diabetes mellitus and daytime sleepiness apnea [13, 14]. At least for workers, sleep problems can lead to serious health problems associated with excessive daytime sleepiness, including hypertension and cerebral vascular disease [15]. Considering Japan’s aging population and falling birth rate, protecting the labor force is imperative by preventing or treating sleep disorders early. The outcomes of this study may contribute to the development of novel approaches to support OD-based sleep disorder prevention or early intervention. In Japan, some reports have suggested a link between tongue pressure and obstructive sleep apnea [16]. We chose to identify and evaluate the OD of workers from oral health examinations because the absence of subjective symptoms raised the possibility that many patients with sleep disorders may go unnoticed. Several restrictions were applied to this study. First, we were unable to examine causal relationships between tongue pressure, lip-seal strength, and OD because this study was based on a cross-sectional survey. Second, there were far fewer females than males in this study; therefore, it was impossible to examine sex differences. Although some research has indicated sex variations in lip-seal strength, the next poll should examine sex disparities in oral health, including OD [17, 18]. Third, in this study, evaluating how the COVID-19 epidemic affected the participants’ dental health-related lifestyle habits was impossible. Changes in the lifestyle (particularly in terms of BMI and eating habits) of workers caused by COVID-19 preventative initiatives may have impacted the measurement of OD [19, 20]. Future longitudinal studies will be necessary to accurately assess the pandemic’s impact [21].

## Conclusions

We found that tongue pressure and lip-seal strength may be associated with OD even in young and middle-aged adults. We believe that measuring OD through dental screening of workers will lead to disease prevention across a wide age range, despite the need to carefully assess the pandemic’s impact.

## Data Availability

Only after acceptance of the manuscript for publication so that we can ensure their inclusion before publication.

## Acknowledgments

We express our deepest gratitude to Okuchy Co., Ltd. and all those who participated in the measurements during the COVID-19 pandemic.

